# A Review of AI and Data Science Support for Cancer Management

**DOI:** 10.1101/2020.08.07.20170191

**Authors:** E. Parimbelli, S. Wilk, R. Cornet, P. Sniatala, K. Sniatala, S.L.C. Glaser, I. Fraterman, A.H Boekhout, M. Ottaviano, M. Peleg

**Affiliations:** University of Pavia; Poznan University of Technology; Amsterdam UMC; Netherlands Cancer Institute; Polytechnic University of Madrid; University of Haifa

**Keywords:** Cancer, Decision Support System, Data Science, Data Integration, Patient Reported Outcomes, Quality of Life, Artificial Intelligence, Predictive modeling, Patient coaching

## Abstract

**Introduction:** Thanks to improvement of care, cancer has become a chronic condition. But due to the toxicity of treatment, the importance of supporting the quality of life (QoL) of cancer patients increases. Monitoring and managing QoL relies on data collected by the patient in his/her home environment, its integration, and its analysis, which supports personalization of cancer management recommendations. We review the state-of-the-art of computerized systems that employ AI and Data Science methods to monitor the health status and provide support to cancer patients managed at home.

**Objective:** Our main objective is to analyze the literature to identify open research challenges that a novel decision support system for cancer patients and clinicians will need to address, point to potential solutions, and provide a list of established best-practices to adopt.

**Methods:** We designed a review study, in compliance with the Preferred Reporting Items for Systematic Reviews and Meta-Analyses (PRISMA) guidelines, analyzing studies retrieved from PubMed related to monitoring cancer patients in their home environments via sensors and self-reporting: what data is collected, what are the techniques used to collect data, semantically integrate it, infer the patient’s state from it and deliver coaching/behavior change interventions.

**Results:** Starting from an initial corpus of 819 unique articles, a total of 180 papers were considered in the full-text analysis and 109 were finally included in the review. Our findings are organized and presented in four main sub-topics consisting of data collection, data integration, predictive modeling and patient coaching.

**Conclusion:** Development of modern decision support systems for cancer needs to utilize best practices like the use of validated electronic questionnaires for quality-of-life assessment, adoption of appropriate information modeling standards supplemented by terminologies/ontologies, adherence to FAIR data principles, external validation, stratification of patients in subgroups for better predictive modeling, and adoption of formal behavior change theories. Open research challenges include supporting emotional and social dimensions of well-being, including PROs in predictive modeling, and providing better customization of behavioral interventions for the specific population of cancer patients.

## 1. Introduction and background

Computerized systems that support clinical decision making and action management often use artificial intelligence (AI) methods to represent, mine, and reason with medical knowledge. These systems demonstrate human-like **artificial intelligence** for supporting medical diagnosis, prevention, and care [1].The knowledge that forms the basis for the AI-based support is acquired from clinical guidelines, evidence-based studies or experts, or it can be mined from electronic health records (EHRs) [2].With the explosion in the availability of health data, collected in EHRs or acquired via wearable sensors, the scope of AI methods in medicine [3] has widened to include such data-centric methods that may be used to generate medical knowledge that drives the AI-based systems.

Data is at the heart of **data science (DS) methods**, which address all stages of the data life cycle [4]. These stages encompass data generation and collection; data processing, integration, storage, and management; data analysis; and finally, data visualization and interpretation; these stages are often grouped into a smaller number of higher-level categories [5]. We grouped them into four categories: data collection (covering data generation and collection); data integration (covering processing, integration, storage, and management); data analysis, and in particular predictive modeling; and results communication (covering visualization and interpretation) - See Figure 1.

**Figure 1.**
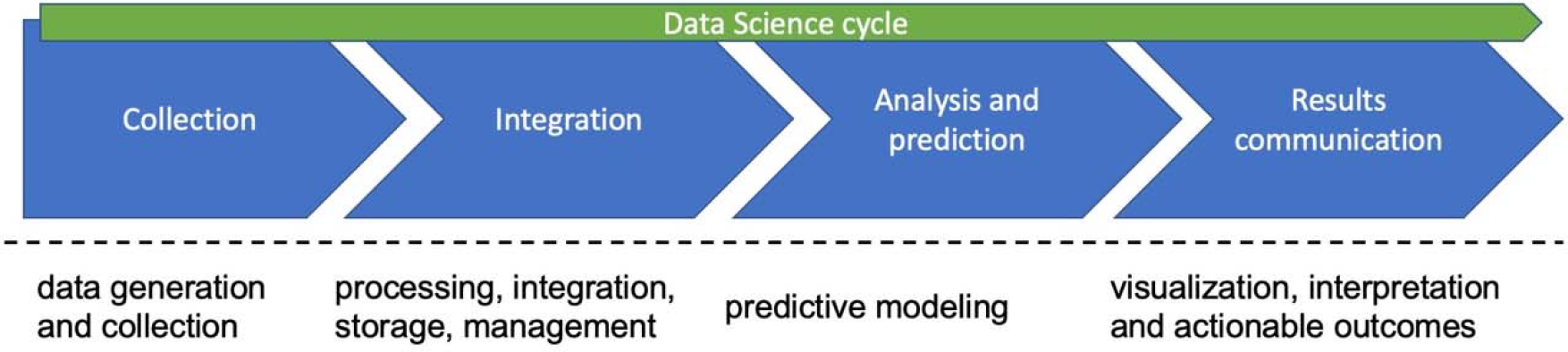
Data Science cycle and categories defining the scope of the literature review. Under the dashed line, mapping of our grouped categories to the taxonomy of phases proposed in [4].

The use of DS methods is already demonstrating their potential for transforming medical care, including radiology[6], surgery [7] and laboratory medicine [8].The focus of this review is on the use of DS for developing AI-based systems supporting patients managed at home and the healthcare providers caring for them. Such a topic has been addressed before in a workshop held in 2017 on innovations in emerging technology, user-centered design, and data analytics for behavioral health [9]. The workshop focused on wearable and mobile technologies being used in clinical and research contexts, with an emphasis on applications in mental health and health behavior change. The workshop included panels on mobile sensing, user experience design, statistics and machine learning, and privacy and security, and led to the identification of research directions for this important and emerging field of applying digital approaches to behavioral health.

Our review focuses in particular on use of DS methods to support care of cancer patients managed at home. After the primary intervention, most cancer patient survivors are managed at home, facing long-term (sequelae) treatments, making the disease comparable to a chronic condition [10]. Despite their benefit, strong therapeutic regimens often cause toxicity, severely impairing quality of life. This may decrease adherence to treatment, thus compromising therapeutic efficacy. Also due to age-related multimorbidity, patients and their caregivers develop emotional, educational and social needs. The CAncer PAtients Better Life Experience Horizon 2020 project (CAPABLE, H2020 875052) will develop a cancer patient coaching system with the objective of facing these needs, in the effort to fully exploit the potential of Data Science to support cancer care and bring it to patients’ homes.

Following the data life cycle approach described above, in this article, we review the literature related to DS methods that cover the entire data life cycle, including: (a) data collection (i.e. monitoring cancer patients in their home environments via sensors and self-reporting);(b) semantic data integration; and based on it provide (c) inference and prediction of the patient’s state; and (d) deliver visualization, interpretation, and communication, which for cancer patients monitored at home, is in the form of coaching, including behavior change interventions.

CAPABLE will indeed use a DS approach in order to provide support to patients and clinicians in two different pathways (Figure1); after (1) collecting data and (2) semantically integrating it, these data can be used to (3) build predictive models that help clinicians infer the patient state and (4) drive patient coaching systems that provide patients with personalized evidence-based recommendations on how to improve their wellbeing. Because the data science cycle encompasses all of these four areas, they define the scope of the review [11]. The findings of the analyses in this review will be used to identify best practices and open research challenges that go beyond state of the art.

## 2. Methods

### 2.1. Search strategy

We designed a review study in compliance with the Preferred Reporting Items for Systematic Reviews and Meta-Analyses (PRISMA) guidelines [12]. We conducted a literature search of the PubMed repository for studies published in the last 5 years (2014-2019) describing AI-enabled systems focusing on the monitoring and decision support of cancer patients. The last search was conducted on March 15, 2020. Our search strategy consisted in the union of four search queries, one addressing each of the identified sub-topics: data collection, data integration, AI/prediction models and coaching systems. The queries were built looking for search terms in TextWord [tw] (which includes title, abstract, Mesh terms and author-identified keywords) in order to increase query recall compared to searching just the title [ti] [13]. Queries included *(system[tw] OR app[tw] OR application[tw])* in order to identify research that produced some sort of computerized clinical decision support system. Finally, specific terms for each of the four sub-topics were added to scope down each of the specific search queries. We report below the query used to collect papers dealing with coaching systems for cancer patients, while the complete search strings employed in the article identification phase are reported in supplementary Table s1. After collecting the results in a Zotero library (www.zotero.org), duplicates were removed. Titles and abstracts of each paper were screened using Abstrackr [14], and irrelevant articles removed before full-text analysis and quality appraisal.

> *cancer[tw]*
>
> *AND (system[tw] OR app[tw] OR application[tw])*
>
> *AND ((coach[TW] OR coaching[tw]) OR (“behavior change”[tw] OR “behavioral change”[tw]))*

### 2.2. Eligibility criteria

We considered articles written in English and published in peer-reviewed journals or conference proceedings in the last five years. Upon initial screening of title and abstract we excluded articles meeting any of the following criteria:

1. Papers dealing exclusively with imaging, genetics or other -omics
2. Papers on statistical survival analysis
3. Papers not dealing with home/outpatients
4. Papers not including the development of a computer-based DSS or computerized behavioral interventions
5. Papers describing protocols for future studies
6. Papers discussing patients being managed for undiagnosed cancer (i.e., no screening tools, even if they are computer-based)
7. Papers on novel drug development, prediction of response to a certain drug, drug-drug interaction, and drug repurposing
8. Papers dealing with animal models (i.e. no human patients)

Note that review articles were excluded from the review itself, but were used to inform the discussion in the present article. Similarly, the references of such review papers were screened to identify relevant articles which, when found, where added to the list of articles considered in the review. The complete list of articles analyzed in this review appears in the Bibliography.

### 2.3. Quality appraisal

All manuscripts included in the present review underwent a formal quality appraisal following the critical appraisal tool from CASP [15]. We selected the CASP qualitative checklist, as it resulted the most widely applicable to the set of articles we reviewed. The results of the quality appraisal have been included in the supplementary material as supplementary Table s2.

## 3. Results

Figure 2 reports the PRISMA flow-diagram of the systematic review. A total of 179 papers were included in the full-text analysis. 70 articles were excluded after accessing their full text, for the reasons listed in Table 1.

**Table 1.**
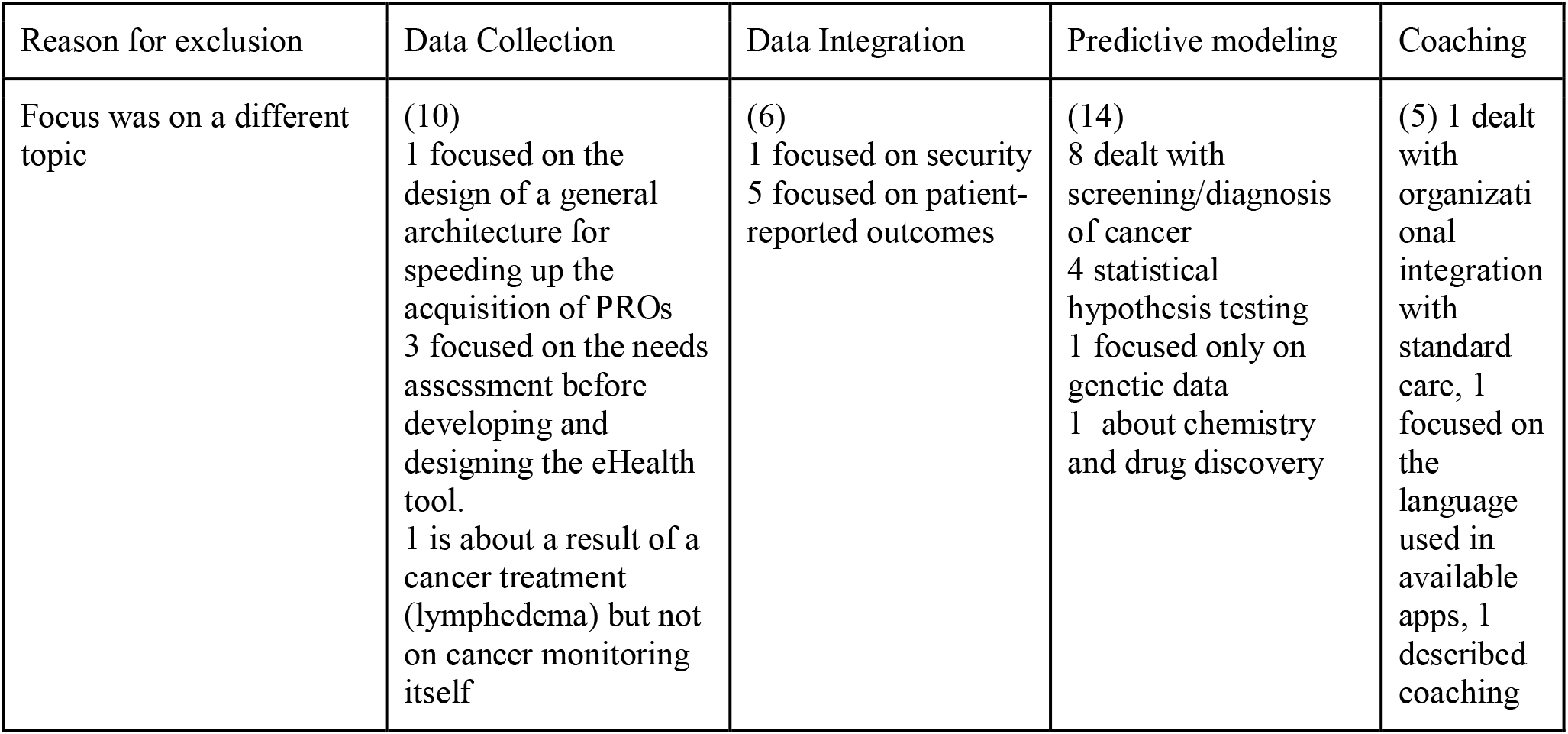
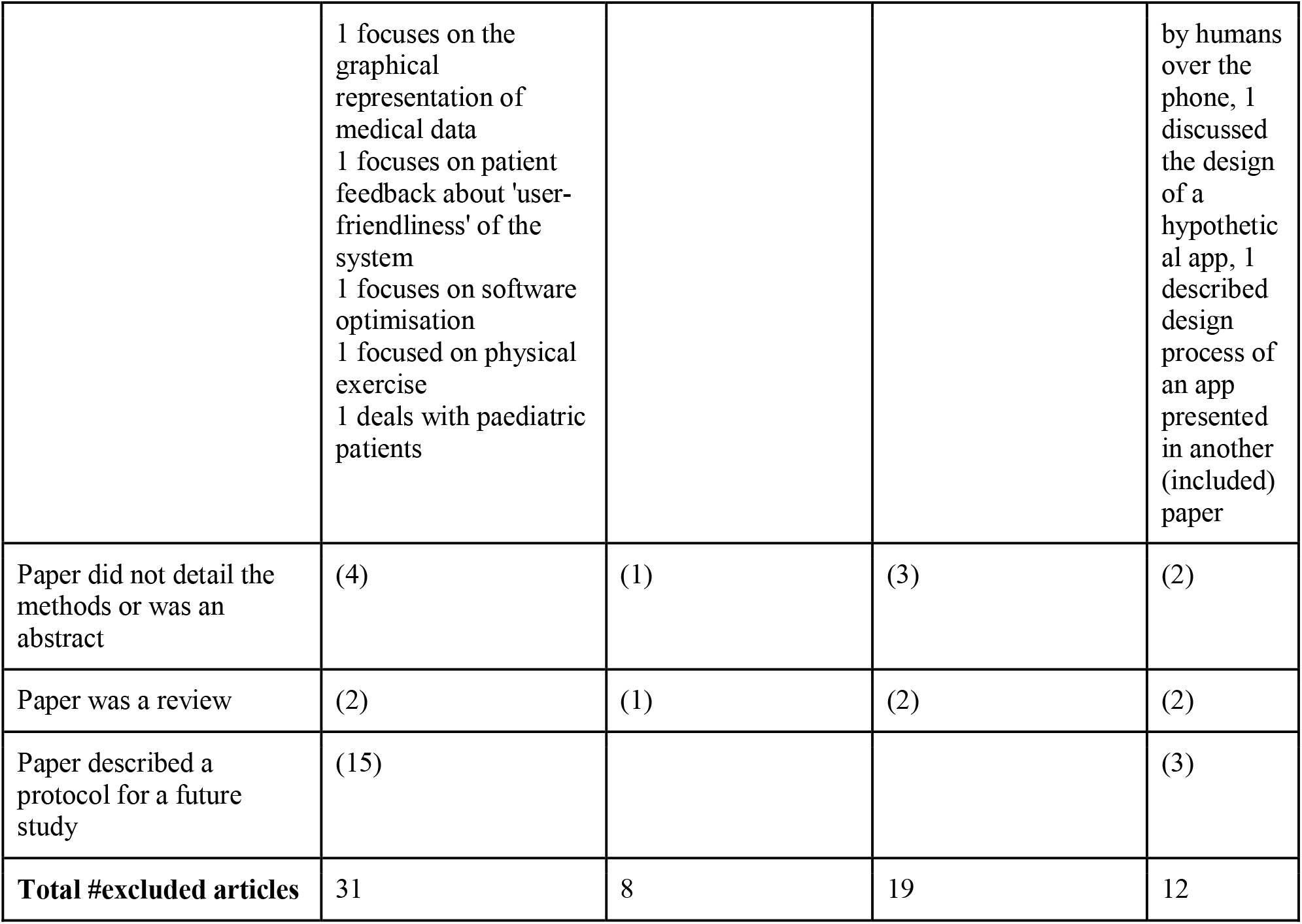
Number (n) of articles that were excluded, after full-text analysis, for each subtopic of the review and the reasons for exclusion.

| Reason for exclusion | Data Collection | Data Integration | Predictive modeling | Coaching |
| --- | --- | --- | --- | --- |
| Focus was on a different topic | (10)<br>1 focused on the design of a general architecture for speeding up the acquisition of PROs<br>3 focused on the needs assessment before developing and designing the eHealth tool.<br>1 is about a result of a cancer treatment (lymphedema) but not on cancer monitoring itself | (6)<br>1 focused on security<br>5 focused on patient-reported outcomes | (14)<br>8 dealt with screening/diagnosis of cancer<br>4 statistical hypothesis testing<br>1 focused only on genetic data<br>1 about chemistry and drug discovery | (5) 1 dealt with organizational integration with standard care, 1 focused on the language used in available apps, 1 described coaching |
|  | 1 focuses on the graphical representation of medical data<br>1 focuses on patient feedback about 'user-friendliness' of the system<br>1 focuses on software optimisation<br>1 focused on physical exercise<br>1 deals with paediatric patients |  |  | by humans over the phone, 1 discussed the design of a hypothetical app, 1 described design process of an app presented in another (included) paper |
| Paper did not detail the methods or was an abstract | (4) | (1) | (3) | (2) |
| Paper was a review | (2) | (1) | (2) | (2) |
| Paper described a protocol for a future study | (15) |  |  | (3) |
| **Total #excluded articles** | 31 | 8 | 19 | 12 |

**Figure 2.**
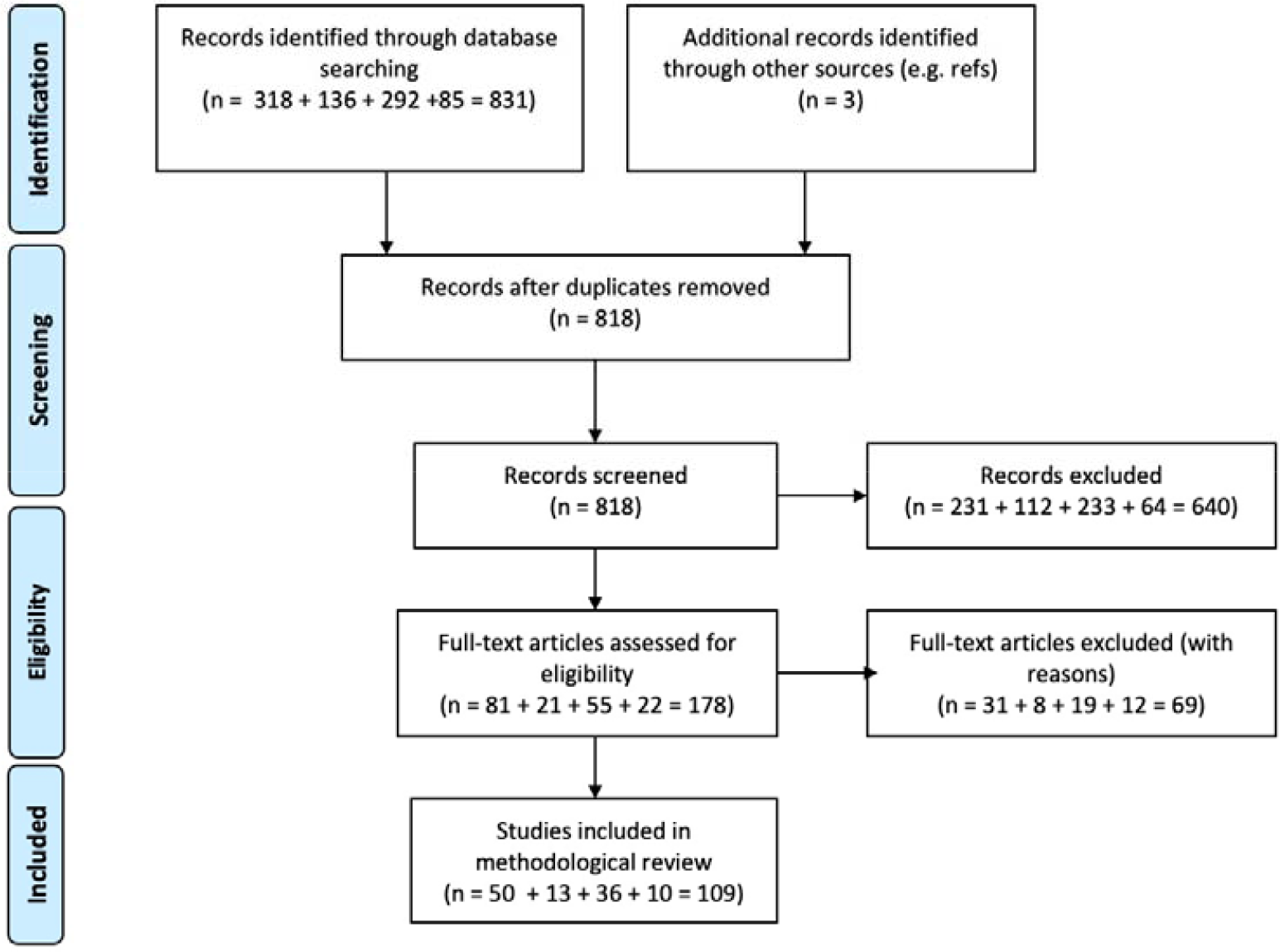
PRISMA flow diagram. The numbers reported at each stage before the total refer to the data collection, data integration, predictive modeling and coaching subtopics.

### 3.1. Data collection

In order to monitor health status following cancer treatment, different types of data should be collected; in addition to physician-provided clinical data, laboratory tests, and imaging results, the data should include patient-provided data and sensor data. Patients may complete patient-reported outcomes (PROs) to report data actively, without an interpretation of patients’ response by a clinician or anyone else. PROs are usually collected through (online) questionnaires. They assess different constructs, consisting of disease or treatment related symptoms, (general) health status or health-related quality of life (HRQoL) outcomes. Another form of collecting data from patients is via sensors. Mobile sensors may be used to collect vital signs from patients in their home environment.

#### 3.1.1. Methods of collecting data from patients

The majority of papers in the Data Collection category (49 of 50) collected data via manual input through electronic PROs implemented inside mobile apps running on smartphones, tablets or using dedicated websites. More popular collection methods were mobile apps, used in 59% of the papers, vs. websites, used in 41% of the papers. In ten papers, sensor data was automatically gathered using sensors embedded in smart bands, smartwatches and other electronic devices. Smartphones were used mainly to gather data from dedicated sensors, which measured physical activities (step count) and vital signs (weight, blood pressure, oxygen saturation, pulse, heart rate and temperature). Among the reviewed papers, “built-in” smartphones’ sensors were not used to collect data.

#### 3.1.2. Patient Reported Outcomes

Linton et al. [16] reviewed 99 self-report measures for assessing well-being in adults and categorized them into six themes; the four dimensions from patients’ life mentioned above, and in addition, spiritual well-being and personal circumstances. According to Linton et al.:

##### Physical well-being

“refers to the quality and performance of bodily functioning, including experiences of pain and comfort.

##### Social well-being

“concerns how well an individual is connected to others in their local and wider social community”.

##### Mental well-being

“assesses the psychological, cognitive and emotional quality of a person’s life”, including the thoughts and feelings that individuals have about the state of their life, and a person’s experience of happiness”.

##### Activities and functioning

“is the behaviour and activities that characterise daily life and our ability to undertake these tasks”.

##### Spiritual well-being

“is concerned with meaning, a connection to something greater than oneself and in some cases faith in a higher power”.

##### Personal circumstances

“are related to the conditions and external pressures that an individual faces, including financial security”.

Patient Reported Outcomes (PROs) are usually collected online, through questionnaires, and assess different constructs, consisting of disease or treatment related symptoms, (general) health status or health-related quality of life (HRQoL) outcomes. HRQoL is fundamentally subjective and multidimensional. HRQoL is measured from the perspective of the patient and covers a range of dimensions from patients’ life, including physical-, social-, emotional-/mental-, and functioning [17]. The European Platform of Cancer Research (EORTC) adds that HRQoL covers the subjective perceptions of the positive and negative aspects of cancer patients’ disease symptoms and side effects of treatment, and stresses the cognitive aspect of functioning [18].

PROs focusing on HRQoL were measured generally before and after the intervention instead of during the use of the eHealth tool, whereas PROs regarding symptom monitoring were collected during the intervention and treatment. Computer Adaptive Testing (CAT) can be applied to shorten the number of questions being asked. CAT is a method to select item sets for individual patients based on a patient’s responses to previous items. The algorithm then selects new items from item banks to maximize the obtained information. Therefore, fewer items are needed to precisely measure patients’ scores. Patient scores can also be directly comparable, even when not answering the same item lists, which is done by item response theory (IRT) methods [19].

In most included papers, combinations of questionnaires were used to collect patient-reported data. Most articles used at least one validated PROs questionnaire and comprised large sample sizes (up to 4345 patients). Patient populations were biased since recruited patients were a-priori in favor of using an eHealth application. PROs questionnaires used for data collection in this review are listed in Table 2. This table presents the reviewed PROs according to well-being dimensions that were defined by Linton et al. [16]. Furthermore, this table presents additional PROs on symptoms. PROs were added to this table when used in 3 or more papers.

**Table 2.**
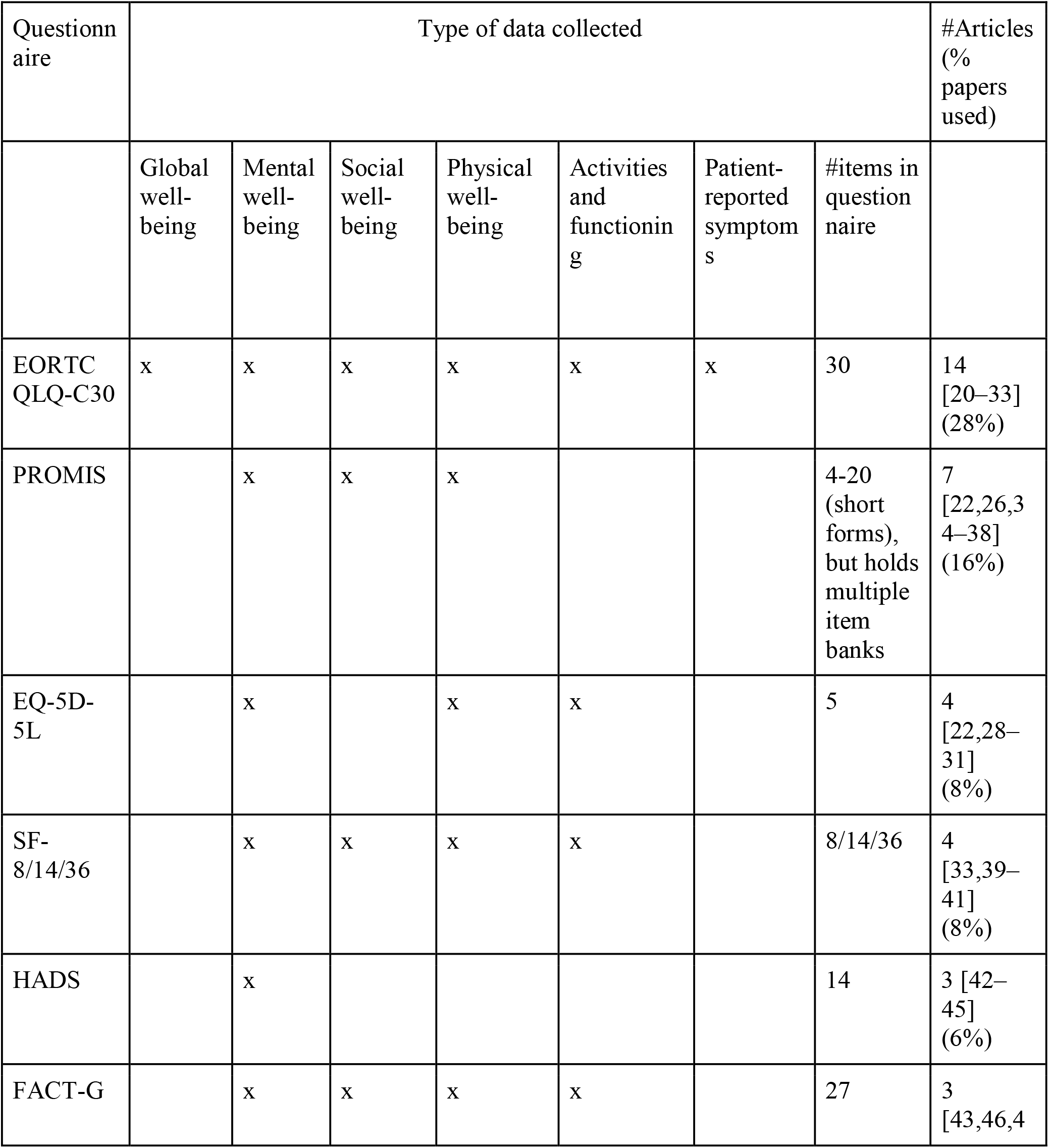
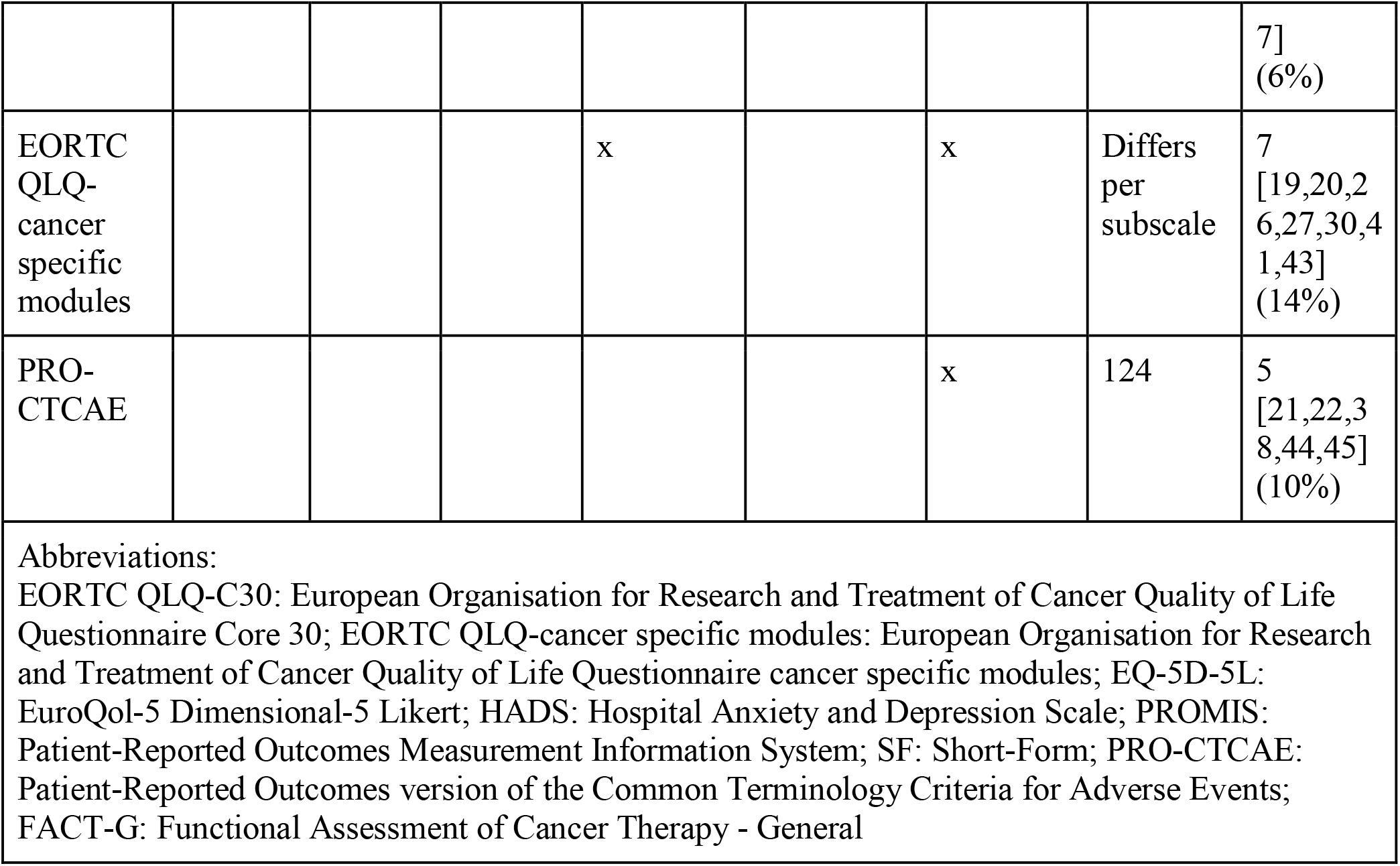
Patient Reported Outcomes (PROs) questionnaires used in the studies. Questionnaire instruments listed in the table are all validated and have been used by at least 3 studies.

##### 3.1.2.1 Patient Reported Outcomes: health-related quality of life

In this review, the most commonly used validated PRO questionnaire for collecting HRQoL data is the EORTC QLQ-C30, used in 28% of the 50 included papers [20–33]. It is a European widely used core generic questionnaire for cancer patients, and comprises most dimensions of well-being as described by Linton et al. [16]l, excluding spiritual wellbeing and personal circumstances. In fact, these dimensions were not captured by any of the PROs used in the reviewed papers. The Functional Assessment of Cancer Therapy - General (FACT-G), also a cancer-specific HRQoL questionnaire is more frequently used in America, but was less frequently used for data collection -- in 6% of included papers [43,46,47]. The second most used questionnaire in this review was Patient-Reported Outcomes Measurement Information System (PROMIS), a set of person-centered measures (enabled by CAT and IRT), used in 16% of the papers [22,26,34–38]. Third most used (8%) questionnaire was the EQ-5D-5L [22,28–31] questionnaire that can be used to compute Quality-adjusted Life Years (QALY’s), a generic measure of disease burden, including both the quality and the quantity of life lived. Therefore, the EQ-5D-5L questionnaire can be used for cost-effectiveness analyses.

##### 3.1.2.2. Patient Reported Outcomes: symptoms

Several PRO questionnaires were used to collect data regarding patient reported symptoms. The EORTC QLQ-cancer specific modules were used to collect cancer-specific patient-reported symptoms in 7 out of 50 included papers [19,20,26,27,30,41,43]. These modules are specific to tumour site, treatment modality, or a HRQoL dimension, to be administered in addition to the core questionnaire (EORTC QLQ-C30) [44]. Furthermore, the Patient-Reported Outcomes version of the Common Terminology Criteria for Adverse Events (PRO-CTCAE) was a commonly used (10%) questionnaire to capture information on symptomatic adverse events [21,22,38,44,45]. The PRO-CTCAE Item Library includes 124 items representing 78 symptomatic toxicities drawn from the Common Terminology Criteria for Adverse Events (CTCAE), reported by patients [48].

#### 3.1.3 Collection of patient data in their home environment using sensors

In this section we discuss the use of sensors to automatically collect additional data (typically vital signs) in the patient home environment. Such data complements PROs captured with questionnaires described in the previous section. Since using sensors imposes no (or limited) burden on the patient (no manual data entry), such modality can be used for frequent and long-term data collection. Reviewed papers described several systems of diversified complexity in terms of the range of employed sensors and the scope of collected data. Group 1 systems consist of a mobile device (smartphone or tablet) connected to a smartband or bracelet (an app usually performs preprocessing and transmits data to a backend; interestingly, none of the described systems relied on sensors embedded in a smartphone). Group 2 is further extended by external sensors (like activity tracker or accelerometers attached to the body), while the smartphone still acts as a hub where the data is collected, processed and transmitted. Group 3 includes systems with standalone devices, which work independently (sometimes) without smartphones. They are more complex and utilize specialized devices like EEG/EMG monitors.

The first group includes simple systems where a smartphone or a tablet is expanded with a wearable device, typically a consumer smartband. Soh et al. [22] described an app providing self-monitoring for gastrointestinal cancer patients. In addition to electronic questionnaires, patients were equipped with wearable bands connected to the app via Bluetooth and used to collect physical activity (PA) information, such as steps count and burned calories. A similar solution was presented by Sun et al [49]. It employed online questionnaires to record HRQoL and smartbands (Vivofit 2 and Garmin Ltd) to monitor PA. PA was monitored by checking the number of patient’s daily steps during recovery. Data from the smartband was being collected 3 to 7 days before surgery, during the patients’ stay at the hospital and approximately 2 weeks after their release from the hospital. Finally, Pavic et al. [32] described a system where patients were provided with a smartphone with preinstalled ‘‘Activity Monitoring’’ app and a commercial sensor-equipped bracelet (Everion). The app recorded motion sensor data (GPS, acceleration), phone call statistics (anonymized), and selected vital signs measured by the bracelet, which was automatically connected to the smartphone via Bluetooth Low Energy. The collected data included step count, step speed and resting heart rate (RHR). Encrypted data was sent once daily to a secured server.

The second group includes systems with additional sensors, which transfer collected data to the smartphones. Timmerman et al. [50] described a system for collecting self-reported symptom severity, mood, medication use and weight. Along with the smartphone, the patient was also equipped with three sensors attached to the body -- an accelerometer (to measure PA). heart rate sensor and oxygen saturation sensor (PPG). Data collected by the wearable devices and the symptom scores were summarized into graphs and made available to doctors and patients. Due to adding wearable sensors, results collected via the symptom questionnaires could be confronted with the collected vital signs. Nápoles et al. [38] presented a solution where a commercial-grade activity tracker was used together with the app to monitor and display progress toward a personalized daily steps goal.

The third group of systems includes solutions where multiple sensors are used in order to collect the required information. Rahman et al. [26] presented a Secure Occupational Therapy (OT) Framework, which aims at HRQoL monitoring. The proposed system uses multiple sensors to detect activities of daily life. The OT movements are mapped to QoL measurements metrics. The collected data can be shared with an oncologist or palliative care unit for real-time decision support. The hardware includes a set of IoT devices such as an LIFX light bulb, door lock, Emotiv EEG device, MYO EMG device, LEAP motion hand gesture tracking sensor, Kinect body gesture tracking sensor, and Eye Tribe pupil tracing sensor. The captured data is transmitted through the secure digital wallet to distributed apps (DApp). These apps apply the privacy and encryption model and offload the data to the edge network for further storage and sharing. Authors proposed DApps for patients, family members, cancer therapists, a secure online doctor on demand, hospital personnel, and medical IoT devices to be able to record the data of cancer patients, caregivers to be able to be in touch with the patients, and data management planning.

We note that using Kinect (and similar) sensors to monitor and provide feedback to patients performing intervention or rehabilitation exercises is an upcoming trend in healthcare. However, the joint positions measured by the Kinect sensor are often unreliable, especially for joints that are occluded by other parts of the body. In addition, users’ motion sequences differ significantly even when doing the same exercise and are not temporally aligned, making the evaluation of the correctness of their movement challenging. Chiang et al. [51] presented a Kinect-based intervention system, which can guide the users to perform the exercises more effectively.

Metcalf et al. [52] presented a health care application to provide more extensive patient education and more thorough perioperative monitoring. Patients with home Wi-Fi access were equipped with tablets preloaded with the health care application (m.Care), an accelerometer and additional equipment for measuring selected vital signs. Specifically, patients participating in the study were provided with the wireless weight scale, the upper arm blood pressure monitor, and the external wireless pulse oximeter. All of these devices were Bluetooth capable and could sync directly to the m.Care application. Participants were asked to perform 5 vitals assessments daily (weight, blood pressure, oxygen saturation, pulse and temperature) for 5 days preoperatively and daily upon hospital discharge until the first postoperative visit.

As can be seen from the above examples, the majority of systems measure the PA of patients. This information is collected based on step count, step speed, which are collected by accelerometers [22,32,50] and burned calories [22]. Solutions that are more complex measure heart rate, oxygen saturation [50] or weight, blood pressure, oxygen saturation, pulse and temperature [52]. There are also Kinect-based solutions, which can help patients to perform the recommended physical exercises [22,51].

### 3.2. Data integration

Data integration is needed when data are partitioned horizontally (i.e., when the same attributes are stored in multiple datasets for different individuals) and/or vertically (i.e., when attributes about individuals are spread out over multiple databases). Several of the reviewed papers highlight the need to integrate data from different organizations and systems to allow creating a big-data cohort, in terms of volume and variety of data. Such cohorts could be analyzed statistically [53], can allow clinical study data to be searched and compared, or can be used to create more accurate predictive models using machine learning methods[53–55]. Other works focus on the need to apply generic models of clinical decision support onto patient data in order to generate patient-specific recommendations [56,57].

Our review focuses on methods for the data integration task. The methods used in the papers that we reviewed leverage patient data standards (information models) or ontologies/terminologies to facilitate the data integration task. Third, integration facilitated by machine-learning methods is addressed.

#### 3.2.1. Data integration via information models

The review paper by Bodenreider [58] stresses that HL7 standards support information exchange by providing an information model for different types of patient data, such as observations, medication requests, procedures, and encounters. Such HL7 classes include attributes for specifying the time stamp, the focus of messages (e.g., (e.g., blood pressure observation, Gemcitabine medication request) according to controlled terminologies such as ICD, LOINC, SNOMED CT and RxNorm, and more specialized attributes, depending on the FHIR class.

Indeed, several of the data integration systems that we have reviewed rely either on simple HL7 messages [59] or on the HL7 Fast Healthcare Interoperability Resources (FHIR) standard [53,56,60] to provide semantics to the data that is integrated and allow standardized access to it. In some works, the databases that are accessed via FHIR-APIs, already provide clinical semantics that are richer than simple storage of medical terminology codes. For example, in KETOS [53] an extract, transform, load (ETL) process exports data from EHRs or data warehouses into an Observational Medical Outcomes Partnership (OMOP) common data model (CDA) database. In the work of Ulrich et al. [61], consortium data is uploaded into an i2b2 (‘Informatics for Integrating Biology and the Bedside’) database. Furthermore, that framework supports the definition of relations between individual data elements and their specification with the help of a suggestion system that uses the LexEVS terminology system of the National Cancer Institute. In [62], the authors develop and leverage a repository of medical data models, represented using CDISC ODM (Clinical Data Interchange Standards Consortium - Operational Data Model https://www.cdisc.org/standards/data-exchange/odm) format.This repository enables manual semantic annotation of data models, as well as construction of data sets that comply to existing data models.

Still, data integration using proprietary information models occurs. For example, Hill-Kayser et al. [57] developed an ETL process to populate LIVESTRONG Survivorship Care Plans with Epic EMR data so that these care plans become patient-specific.

Some systems also use other standards. For example, [56] describes instantiation of a Bayesian Network for a patient using a combination of Arden Syntax Medical Logic Modules that specify the logic of the required processing of data. The “curly brace” statements in Arden Syntax describe the needed data elements by calling a FHIR resource that refers to the patient ID or a SNOMED code for clinical findings. Ulrich et al. [61] use the ISO 11179-3 metadata repository standard. Metadata about data elements is represented according to the FHIR standard. For example, Case Report Forms are represented as a FHIR questionnaire. The FHIR-based processing allows exchange of data elements with clinical and research IT systems as well as with other metadata systems.

##### 3.2.1.1 Integration of genomics and EHR data

While the integration challenge often involves reaching consensus on similar data elements represented differently, either in different information models, or using different vocabularies, an additional challenge is the integration of vertically partitioned data of different modalities, coming from different realms, such as genomic data and clinical data. Several of the reviewed papers discuss this emerging need for integrating genomic (omics) data and clinical and epidemiological (non-omics) data. Integration of omics and non-omics (OnO) data shows promise in improving predictive models, phenotyping patients and finding clinical risk factors. Integration of cancer genomic data with EHR systems could also improve clinical decision making and the use of personalized care. However, efforts are still scarce as integration of OnO data poses a range of challenges, such as the heterogeneous nature of clinical data and the lack of consistent standards for both genomic and clinical data. López de Maturana et al. [63] reviewed attempts of OnO data integration in clinical and epidemiological studies. Although few of their reviewed studies explicitly address this type of modeling, joint modeling approaches are recommended by López de Maturana et al. [63] for integration of large-scale OnO data. Joint modeling approaches capture a larger complexity than the modeling approaches more frequently seen such as conditional or independent modeling. Additionally, joint modeling accounts for the correlation structure between the two data types.

Mihaylov et al. [64] present a framework for this joint modeling of clinical and omics data. A network is presented with the aim to predict survival time. Utilizing semi-structures for each data type (such as clinical, expression and mutation data), relations are found that represent the internal network. Enrichment of these data and relations is possible by linking data to external domain knowledge sources. Mihaylov et al. show the utility of semantic OnO data integration as survival time prediction models bear improved results using the relational OnO data network compared to separate omics and non-omics survival prediction.

Focusing on integrating genomic data with EHR systems, Warner et al. [65] discuss the status (as of 2016) and the challenges. Although well-established nomenclatures exist for omics data, a lack of consistent modeling standards prevents integration of omics laboratory results into EHRs. Currently, laboratory results are included in PDF format which does not allow for any secondary use of data or for clinical decision support. Deemed as most useful of the emergent solutions is the use of application programming interfaces (APIs) or RESTful web services. Particularly, SMART on FHIR clinico-genomics apps were highlighted as solutions that incorporate the presentation and contextualization of genetic test results into the clinician workflow.

#### 3.2.2. Data integration via ontologies and vocabularies

The review paper by Bodenreider [58] provides an overview of how ontologies can be used for data integration. First, in the warehousing approaches, controlled vocabularies can be used to describe the focus of data items in local sites, facilitating their later integration. The integration can span data sets with different content that may be combined. For example, LOINC has been used for integrating laboratory data with adverse events, the Foundational Model of Anatomy for the integration of genomic information sources, and SNOMED CT for the integration of disease and pathway information.

Second, mediation-based approaches use ontologies for defining a common global schema for data elements (common data elements, CDE) and mappings between the global schema and local schemas are then defined. This allows composing queries to the local schemas in terms of the common schema.

The data integration approaches from recent years use a combination of both approaches. Several papers [53,54,61,66–68] present ETL database functions that allow pulling data from a source database and placing it into a target database. The first three of these papers define manual processes by which local sites define mappings between source data elements (SDE) to a set of needed common data elements (CDE). Use of controlled vocabularies such as SNOMED CT and LOINC facilitates the agreement. Similar to the terminology-suggestion system of Ulrich et al [61] described in Section 3.2.1, Mate et al [68] support term matching further via an algorithm that supports fuzzy matching of terms, utilization of synonyms, and sentiment tagging to suggest mapping of SDEs to CDEs. A final step of data alignment replaces the source value sets with those from the target terminology, as well as converts between different data types according to expert-curated mapping rules.

#### 3.2.3 Data integration supported by machine learning

While the methods described above have a focus on manual or rule-based integration based on ETL-procedures, increasingly machine learning methods are applied for data integration. Mariette et al. [55] propose an approach that uses combined kernels in kernel self-organizing maps (KSOM) to cluster and visualize multi-omics breast cancer datasets. This clustering is relevant to discriminate between different breast cancer subtypes and to identify their relations and has the strength that it can be performed in an unsupervised manner, removing much of the burden of integration based on information models and ontologies.

Isoviita et al. [54] present an open-source, cloud-based machine learning system where datasets from multiple (live) sources (EHR databases and research databases) are integrated using extract, transform, load (ETL) processes and melded into a single database, but with minimal transformations. These merged but heterogeneous data are used for the training of ML predictive models. In this case study where primary therapy outcomes in high-grade serous ovarian cancer were predicted, results again demonstrated the benefit of combining information from multiple sources.

These papers show that machine learning enables analyses on integrated data without the need to perform supervised transformation of data, which is generally a resource-intensive task.

### 3.3. Predictive modeling

#### 3.3.1 Input data for predictive modeling

One of the main preconditions for building an accurate and reliable prediction model is the availability of high-quality, large-enough dataset to serve as the model training set. This is true in cancer, as well as in other medical domains. In addition to the choice of a particular ML algorithm, the performance of a predictive model largely depends on what input data has been used for training. The availability, promoted in recent years, of publicly accessible databases and registries focusing on cancer patients has proven to be a major driver of interest for studies on predictive modeling of cancer outcomes. One outstanding example is Surveillance, Epidemiology, and End Results (SEER) database, which has been used in a number of the analyzed articles [69–75]. On the other hand, availability of one own organization original data (e.g. from EHR) can constitute a significant competitive advantage with the possibility of accessing a wider array of patient data variables, and combining them for improved knowledge discovery and ultimately better predictive performance for specific populations. For example Soguero-Ruiz and colleagues [76] were able to leverage EHR clinical notes to build a model to predict anastomosis leakage in patients with colorectal cancer starting from NLP-extracted features. In another study [77] the same authors complemented EHR free text data with detailed labs and vital signs data to improve adverse event prediction. Again, Alabi et al. [78,79] used data coming from a federation of hospitals (in Finland and Brazil respectively) to build prognostic models for oral cancer tumor recurrences.

Large, publicly available datasets can provide the optimal conditions when size of the training data, standardization of variables and reproducibility (also comparability to other published research) of results are considered of the utmost importance. However, dealing with an original, unpublished dataset, usually owned by partnering clinical organizations, can provide a competitive edge in originality of the research and access to an extended number of independent predictors (e.g. detailed labs, or -omics data) that might prove useful for achieving the best predictive performance.

An additional finding that emerged from the analysis of the articles is that current research in predictive modeling should strive to generate generalizable models (i.e. models that perform well when applied to cohorts different from the one used for the original development and validation). Thus the use of a restricted set of easily collectable predictors is to be regarded as a desirable feature. One example can be found in Oliveira et al. [75] where a set of only 6 features from the SEER dataset proved effective in predicting survival of colorectal cancer patients at different time points (year 1,2,3,4 and 5). A notable characteristic of such model is the use of a dynamic set of predictors that evolve over time, to achieve best performance. The fact that the knowledge elicited by ML models should be validated over time is also a focal point to promote model generalizability, especially after some years from the original model elicitation. Indeed Kleinen and colleagues [72] advocate for knowledge embedded in predictive models for breast cancer to be updated every ten years to maintain good performance.

#### 3.3.2 The quest for more complex machine learning models: ensembles, deep learning and the like

Once the issue of possessing a good input dataset to train a well-performing model is solved, the next question researchers in this field face is what ML algorithm to use. In recent years, in light of the big-data era, there has been some debate [80–82] on whether cutting-edge ML models could deliver better performance than well-established statistical methods (e.g. Cox regression for survival analysis) that have been traditionally employed in cancer outcomes research. Whereas most of the articles analyzed employ a quite traditional approach where an array of ML algorithms is applied looking for the best-performing one, some notable exceptions occur. Deep Learning (DL) is for sure one of the paradigms that is drawing attention especially in cases where large and unstructured (e.g. -omics) data is available. The application of DL to medical predictive modeling is out of the scope of the present review (which focuses on predictive modeling mainly using clinical data) and deserves the undivided attention of the research community [83–85]. However we did find two instances where DL was applied within the scope of the present review[86,87]. Another family of models that consistently perform well in cancer outcomes prediction is the one comprising different forms of ensembles: i.e. instances where a set of models is trained, instead of a single one, and their output is combined in various ways in order to provide the final prediction. Some of the most notable are approaches using bagging like random forests (RF)[75,86,88], approaches based on boosting (e.g. XGB or gradient boosted decision trees)[73,89,90] and voting ensembles[71,75]. A rather original approach is employed by Morino [91] who used temporal expert advice (TEA) where a set of “experts”, each represented by another patients’ time series, were combined in a weighted voting scheme. Despite the good performance that more complex ML algorithms provide, it is also worth mentioning the fact that simpler models also bring value, especially when the predictive model has to be deployed in the clinical setting to assist decision making. Complex models indeed quickly lose appeal when interpretability, and ease of use come into the picture. Models where a limited set of features is used as input (e.g. and selection is performed through features selection) may favor implementation in web [78,92] and mobile [75] tools that are ready to use at the bedside. Finally, feature selection and feature engineering can be effective tools to take advantage of prior knowledge, in order to boost an ML model performance [93,94] complementing a purely data-driven approach with expert knowledge.

#### 3.3.3 Precision medicine principles help predictive modeling

Some research efforts about predictive modeling in cancer can significantly benefit from principles promoted by precision medicine, especially considering its strong focus on oncological applications [95]. Subgroup analysis based on patient similarity, clustering based on big data and unsupervised phenotype discovery [96] can all improve cancer outcome prediction.

In the current review, two papers [69,74] highlight how developing stage-specific models, despite the potential reduction in the size of patient-cohorts, proved to be beneficial to performance, while evaluating predictive models for survivability on all the stages together might lead to systematically overestimating their predictive performance.

As in the above example, some studies found evidence that being able to stratify patients before the actual modeling is beneficial. Kawakami et al. [89] advocate that use of predictive algorithms (in particular a combination of supervised and unsupervised approaches) may facilitate personalizing treatment options through pretreatment stratification of patients. Another example of successful application of unsupervised ML algorithms can be found in Lynch et al. [73], who used unsupervised data analysis techniques like self-ordering maps and k-Means to classify patients by defining the classes as effective proxies for survival prediction. Their study on lung cancer patients from SEER suggests comparable results to state-of-the-art supervised regression techniques, such as gradient-boosting machine. Despite some evidence of their potential usefulness, we report an under-utilization of unsupervised ML approaches in the analyzed articles.

### 3.4. Coaching

Given improved cancer treatments and increasing survival rates, there arises a need to manage HRQoL of cancer patients [97]. Evidence suggests that HRQoL is a significant prognostic predictor, where lower levels are associated with poorer survival, yet it is affected by a number of disease- and treatment-related side effects. Multiple attempts of addressing these side effects with digital interventions implemented as mobile or web-based apps have been described in the literature, and in this section we review selected works. In Section 3.4.1 we focus on different aspects of patient behavior that correspond to specific HRQoL dimensions. Moreover, in Section 3.4.2 we present behavior change techniques and other theoretical models employed by the reviewed systems.

#### 3.4.1. Addressed aspects of patient behavior

The prevalent aspect of patient behavior is physical activity (PA) that is discussed in [98–101]. Reviewed studies involve both generic apps developed for general audiences, as well as specialized apps aimed specifically at cancer patients. Use of generic apps and activity trackers for encouraging and monitoring physical activity in breast, prostate and colorectal cancer patients was described in and [98] and [99]. While these interventions resulted in increased physical activity and better awareness of lifestyle (e.g, through idle alerts), the patients pointed at several shortcomings. The major one was the lack of customization for specific patients that would take into account limitations imposed by their disease and treatment. Such customization should for example cover the type of suggested activities with walking being considered as the easiest and safest one, and the frequency and tone of reminders. Other shortcomings included inability to define goals for such activities as swimming or cycling and to automatically track them. Moreover, patients suggested such interventions should be prescribed as a routine part of care and there should be a possibility of discussing them with a healthcare provider, e.g., a nurse specialist.

A dedicated web-based system - RiseTx - for reducing sedentary behavior in prostate cancer patients was described by Trinh et al. [100]. RiseTx offers a multi-phase intervention that involves self-monitoring to establish baseline, action planning and progressive release of self-regulatory strategies, consolidation and maintenance. The system also uses incentives to increase engagement -- patients obtain reward points that they can redeem for various items or donate to charity. In the feasibility study the authors observed significant decrease of sedentary time at post-intervention, but this change was lost at follow-up which suggests challenges with sustaining engagement once the intervention had been completed. Another specialized app targeting PA is BENECA presented in [101] and implemented as a mobile app. BENECA supports cancer patients in managing their energy by monitoring its intake (diet) and expenditure (PA) and by providing immediate feedback and recommendations based on guidelines and systematic reviews. The system was verified in 8-week study - while its use posed some challenges with collecting diet-related data, it improved several QoL scores (captured with EORTC QLQ-C30), e.g., global health, physical functioning or cognitive functioning. Moreover, its users felt more motivated to increase the level of PA.

Other frequently discussed aspects of behavior are related to mental well-being and include depression [102], distress and anxiety [1,2]. In [102] Chow et al. present the iCanThrive mobile app that helps women’s cancer survivors reduce symptoms of depression. The app offers 8 modules with interactive exercises, e.g., challenging a negative thought. In a 6-week pilot study the authors observed a significant reduction in symptoms of depression at post-intervention. Moreover, there was no significant difference between post-intervention and follow-up. Smith et al. in [103] described application of the PTSD Coach mobile app developed by the US Department of Veteran Affairs to manage post-traumatic stress disorder in cancer patients. The app provides educational resources, self-assessment tools extended with interpretative feedback and a set of mind-body exercises. PTSD Coach was tested in an 8-week pilot study where most of the participants were satisfied with the app and nearly half of them reported improvement in their symptoms. Finally, Kubo et al. [104] described the use of the Headspace app to improve mental well-being of cancer patients undergoing chemotherapy and their caregivers. Results of an 8-week study showed benefits in terms of statistically significant reduction of distress, anxiety and depression. It also revealed several interesting findings -- the baseline level of distress and anxiety was higher for caregivers than for patients, and patients who participated with their caregivers demonstrated better adherence with the app and experienced stronger reduction in depression.

The last group of papers focuses on efficacy (improved skills) for handling problems associated with disease and its symptoms [105,106]. Wklander et al. [105] describe the web app developed as part of the Fex-Can (Fertility and Sexuality Following Cancer) project aimed at adolescent and young adults. The app supports the patients in improving their skills to target sexual and fertility-related problems by delivering a mixed educational content including articles, exercises and video vignettes (real stories patients can relate to). The app underwent a 2-month feasibility study where more than half participants remained active throughout that period and considered the content to be relevant and informative. Finally, Beck et al. [106] describe the SymptomCare@Home system focused on patients receiving chemotherapy. The system facilitates reporting of relevant symptoms through IVR (interactive voice response), it also delivers immediate feedback and self-management coaching appropriate for the patient’s state and treatment toxicity. The coaching is evidence-based -- it builds on clinical guidelines and literature and covers a whole range of topics, such as preventing weight gain, improving eating, improving concentration and thinking and relieving pain. The system underwent a randomized cliinical trial where its users had significantly less overall symptom severity and reduction in the number of days with moderate and severe symptoms.

#### 3.4.2. Applied behavior change techniques and other models

Authors of selected studies also discussed behavior change techniques (BCTs) and other theoretical constructs employed in considered apps. First, Kalke et al. [107] reviewed 30 apps for breast cancer patients available to the general public in popular app stores and covering nearly all cancer continuum with the exception of end-of-life support. The authors used the taxonomy for coding BCTs in mobile cancer apps [108] (this taxonomy is based on the original proposal by Abraham and Michie [109]) and identified 12 BCTs coming from 6 categories: customization, information/behavior relationship, intention, facilitation, self-efficacy and social influence. BCTs from the first four categories were the most prevalent ones -- they were found in at least half of the reviewed apps. Moreover, BCTs aimed at facilitation (i.e., providing instructions, materials and education) were implemented in 80% of the apps. The authors also found out that the apps with user star ratings offered significantly more BCTs than these without ratings.

Similarly, Roberts et al. [99] identified BCTs in generic PA mobile apps that can be used by cancer patients. They identified the same categories of BCTs and in the previous study with the exception of customization, however, this difference could be attributed to using the original taxonomy by Abraham and Michie that does not explicitly mention the customization and personalization category. BCTs were also mentioned in the context of the RiseTx system [100] where in addition to intention-related BCTs as action and coping planning the authors also employed contingent awards (incentives) to increase engagement with the intervention. Finally, the BENECA system [101] employed BCTs from the intention, facilitation and self-efficacy categories.

Finally, the authors of the iCanThrive app [102] employed the efficiency model of support that identifies failures that prevent users from benefiting from a digital intervention, including issues related to the usability of the app, its fit to the user’s needs, knowledge on how to use the app and implementation failures. The model was used to develop protocols supporting phone calls aimed at improving engagement with the app, promoting knowledge of the skills found in the app and encouraging implementation of these skills in daily life.

## 4. Discussion

Our analysis of the papers identified dimensions that emerged and were common to papers from the four scope areas. Table 3 summarizes the how papers in the four scope areas addressed the dimensions of cancer type, cancer stage, the evaluation done, and year of publication.

**Table 3.** Common metadata about the papers reviewed. Cancer types have been ordered top-to-bottom according to their incidence (incidence source data [102]).

|  | Scope area | Data Collection<br>(50 papers) | Data Integration<br>(13 papers) | Predictive modeling<br>(36 papers) | Coaching<br>(10 papers) |
| --- | --- | --- | --- | --- | --- |

| Dimension |  |  |  |  |  |
| --- | --- | --- | --- | --- | --- |
| Cancer type | Lung | 3<br>[35,110,111] | 1 [59] | 6<br>[64,66,68,82,83,112] |  |
|  | Breast | 5<br>[24,25,27,38,113] | 5<br>[53,55,57,64,67] | 5<br>[69,72,90,114,115] | 8<br>[72,98,99,101–105] |
|  | Colorectal | 2 [22,47] | 4<br>[53,57,61,68] | 6<br>[26,76,77,90,116,117] | 1 [99] |
|  | Prostate | 2 [18,118] |  | 5 [86,89,119–121] | 3 [93,95,98] |
|  | Head & Neck | 5<br>[29,33,111,112,122] | 1 [56] | 5<br>[78,79,118,119,123] |  |
|  | Gynaecologic | 3<br>[20,41,124] | 1 [50] | 3 [81,84,123] | 1 [97] |
|  | Bladder | 1 [105] |  | 1 [106] |  |
|  | Hematologic |  |  |  | 3 [103–105] |
|  | Other / not specified | 29<br>[20,26,28,30–32,34,37,39,40,43,45,46,49,110,120,121,124–135] | 4 [60,62–64] | 5<br>[69,92,93,136,137] | 3 [104–107] |
| Cancer stage | Diagnosis |  | 1 [55] |  |  |
|  | Treatment | 33<br>[21,21,25,26,28,29,34,39–43,46,49,52,110,111,113,120–122,124,125,127– | 1 [54] | 7<br>[78,91,93,114,119,137,139] | 3<br>[100,104,106] |
|  |  | 135,138] |  |  |  |
|  | Post-treatment | 4<br>[20,23,47,112] | 1 [64] | 5<br>[79,88,90,94,140] | 4<br>[27,98,99,102] |
|  | Both treatment and post-treatment | 3<br>[33,45,50] | 1 [57] | 9<br>[70,76,87,89,92,116,118,123,141] |  |
|  | Palliative | 4<br>[22,30,32,37] |  |  |  |
|  | Other / not specified | 6<br>[24,27,31,35,38,126] | 5<br>[54,61,64,67,68] | 15 [69,70,72–76,78,87,111,115,137,142–144] | 3<br>[104,106,108] |
| Evaluation done | Data set size used for evaluation of integration: |  |  | Applicable only to studies reporting external validation |  |
|  | Small (<100 patients, <50 elements): | Small: 27<br>[21,22,24–26,28,30,31,34,35,37,41,43,47–49,51,112,114,121,123,124,127,135,136,136,138,139] | Small: 1 [54] | Small: 3<br>[79,80,145] | Small: 8 [99–106] |
|  | Medium (<1000 patients, <100 elements): | Medium: 7<br>[23,29,39,40,44,46,110] | Medium: 5<br>[55,56,58,61,66] |  | Medium: 1<br>[107] |
|  | Large: | Large: 3<br>[32,131,132] | Large: 4<br>[57,63,66,67] |  |  |
| Publication year | 2015 | 2 | 1 | 2 | 0 |
|  | 2016 | 3 | 5 | 4 | 0 |
|  | 2017 | 7 | 0 | 9 | 2 |
|  | 2018 | 5 | 2 | 4 | 3 |
|  | 2019 | 17 | 5 | 15 | 3 |
|  | 2020 | 16 | 0 | 2 | 2 |

The above Table 3 allows some interesting observations. First, Breast cancer is the most represented in the group of papers we analyzed. Neither melanoma or kidney cancer, which are the focus of the CAPABLE research project, have been explicitly addressed in any of the retrieved articles. Number of articles focusing on a certain cancer type are correlated to different cancer types incidence. The topic of our review that covered the least prevalent types of cancers is coaching. However lung and head&neck cancers had no coaching system explicitly addressing them, probably because of their short course and limited survival.

Secondly, we chose to only focus on already diagnosed cancer patients. For this reason Table 3 reports cancer stages other than diagnosis, thus starting from the treatment stage. What is more, cancer stage did not make a difference for works about data integration. Finally, the palliative stage is generally under-supported, and indeed no data integration, predictive modeling or coaching paper addressed it. However, it is worth mentioning that, especially for coaching systems, not only patients but also home caregivers are likely to benefit from support in this end-of-life stage.

For what regards size of the evaluation, most papers 37/50 of the data collection area performed some evaluation: most of them (27) on a small sample size, and less (7 and 3) on a medium or large sample. On the other hand, only three articles on predictive modeling performed external validation on small sets of patients. Most (9/10) coaching system papers performed evaluation, albeit on small cohorts.

Finally, a trend for a generalized growing interest in all data science areas can be pointed out from Table 3 data. In particular, electronic data collection and predictive modeling are becoming more popular in the last 2 years.

### 4.1 Best-practices and open research challenges

We present the best practices and open challenges that we identified in each scope area.

#### 4.1.1 Data Collection Best Practices

We identified the following best practices regarding data collection:

a. Use validated questionnaires to collect PROs
b. Choose a set of PRO questionnaires that covers multiple aspects of wellbeing, including physical, mental, social, activities and functioning, and symptoms.
c. EORTC QLQ-C30 has good coverage of well-being dimensions and is validated. It is the most widely used PRO questionnaire in Europe (used in 14 of the 50 studies reviewed in the area of data collection). Specific cancer versions are available or currently under validation (e.g. QLQ-MEL38)
d. Collect outcome data at least at baseline and after the intervention, but preferably during the intervention as well.
e. Presently, sensors offered by smartphones are not a reliable and accurate source of measurable data.

#### 4.1.2 Data Collection Open Challenges

i. Develop easy methods for collecting PROs and symptoms from recovering patients and survivors in their home environments, while they are undergoing the intervention. This could be facilitated by the two methods below.
ii. Develop computer-adapted testing versions of validated PRO questionnaires that have over twenty questions in order to make it easier for patients to complete them.
iii. Reliable and easy to use mobile/wearable sensors could be used to collect vital signs, symptoms and PRO data (e.g., activity and functioning) when possible, relieving patients from the burden of self-reporting of symptoms and from the bias of reporting untruthful data. The degree of correlation between monitors routinely used in clinical practice and the smartphone-based applications is insufficient to recommend clinical utilization of the latter. This lack of correlation suggests that the smartphones-based applications do not provide clinically meaningful data. The inaccurate data provided by these applications can potentially contribute to patient harm.

#### 4.1.3 Data Integration Best Practices

We identified the following best practices regarding data integration:

a. Semantic data integration makes the data set more comprehensive -- larger data sets or data sets that include more data items about the patients. This improves the ability to infer more valid results from data analysis.
b. Data integration should include extract, transform, load (ETL) functions that allow pulling data from a source database (e.g., hospital information system) and storing it in a target database.
c. The ETL process can benefit from basing the target database on the Observational Medical Outcomes Partnership (OMOP) common data model (CDM), as in KETOS [53]. OMOP provides services (based on R) to perform distributed analysis of data based on the OMOP semantics. It will help researchers to compare their data and results. Observational Health Data Sciences and Informatics (OHDSI), which is the organization that develops OMOP, focuses on horizontal partitioning of data sets with similar attributes. Projects that span multiple sites, such as CAPABLE, can benefit from this platform for distributed analysis.
d. Data should meet principles of findability, accessibility, interoperability, and reusability (FAIR). OMOP provides the basis for Interoperability of the data. Findability and Accessibility are facilitated by depositing metadata in a repository (e.g., Zenodo) that describes the data and the access conditions. Reusability is realized by clear and as-open-as-possible licensing, targeting at CC-BY (which allows others to distribute in altered or unaltered form, also commercially, as long as credit is given) and CC-0 (i.e., “No Rights Reserved”).
e. The most popular patient information model used was FHIR. It is a standard used by commercial vendors for health information exchange. Supporting it will ease data exchange and potential commercialization.
f. Standard vocabularies (ICD, LOINC, SNOMED CT and RxNorm) are used by most of the papers to capture the foci of FHIR or OMOP objects.
g. The ETL process can be done manually in a process by which local sites define mappings between source data elements (SDE) to a set of needed common data elements (CDE). The manual process could best be supported by terminology services to locate standard terms and their meaning.
h. The integration process may also be supported by the use of machine learning, either to harmonize data, or to perform analyses on pluriform data that has not been harmonized.

#### 4.1.4 Data Integration Open Challenges

i. Best-practice ETL processes benefit by exporting data from EHRs or data warehouses into an OMOP common data model database. However, OMOP does not provide a data exchange/communication standard. We recommend adding a FHIR API layer to allow components of a complex personalized knowledge-based and prediction-based decision-support system to semantically share data and knowledge through a standardized API. Combining OMOP, FHIR, and controlled terminologies for the ETL target database, leverages the benefits of all of these standards resulting in a “best of breed” approach for storage and exchange.
ii. The papers reviewed above do not address the semantic gap that exists between formalized clinical knowledge used for decision support and raw data stored in integrated ETL target databases. The knowledge sources, such as computer-interpretable clinical guidelines [144], contain abstract terms (e.g., complicated diarrhea) that can be computed from raw data about symptoms that are stored in the database. A component such as Knowledge-Data Ontology Mapper [146] or the Medical Database Adaptor (MEIDA) [147] could be added to bridge the semantic gap by allowing modelers to provide ontological definitions of abstractions in terms of a standard patient data model and based on it, automatic query generation. However, both of these mediators have shortcomings, described in [146], and should be extended to support the full range of abstractions needed for decision support.

#### 4.1.5 Predictive modeling Best Practices

We identified the following best practices regarding predictive modeling:

a. Some articles deal with unbalanced data and cope with it mostly with undersampling.
b. Stratifying patients in subgroups before predictive modeling promotes better predictive performances.

#### 4.1.6 Predictive modeling Open Challenges

i. Predictive models should be validated on unseen patient datasets. Most studies do not perform external validation, but instead rely on more conventional cross validation or train-test splits, which tend to overestimate performance [142,143].
ii. None of the predictive modeling papers have considered using PRO data as predictors of good/bad outcome or survival. The validity of AI-based inference of patients’ psychobehavioral dimensions [148] (e.g., stress, anxiety, sleep quality, depression) from PRO and sensor data needs to be assessed.
iii. Unsupervised approaches are underutilized for prediction of a patient’s state based on clinical, PRO and sensor data.

#### 4.1.7 Coaching Systems Best Practices

We identified the following best practices regarding patient coaching.

a. Behavioral interventions (e.g., physical exercise) for cancer patients offered by apps should be suited for the characteristics of cancer patients rather than be general-purpose. Robertson et al. [149] presented a hypothetical app that provided interesting suggestions given by cancer patients which should be considered:
  - Messages to patients should use a casual (not clinical), concise, and positive tone. This is in line with recommendations from the IOM on the language that should be used in mHealth apps [150]
  - The app should include tools that support personal goal attainment (e.g., reminders, role model narratives)
  - Value-based goals should be included (e.g., “I want to see my grandchildren married”). They should be attained by short-terms goals (e.g., for physical activity) that are easy to attain.
  - Recommendations should come from a trusted source, and should include layman summary of the relevant literature justifying the recommendation
  - Recommendations should preferably include video demonstrations, preferred over text and pictures. Exercises could be presented by other cancer patients.
  - Interventions should deliver an experience that is tailored to the user’s characteristics (cancer-related information, age,…), also taking into account the location of the patient and weather
  - Whenever possible, wearables should be used to track patients; this lowers the burden of data entry.
  - Adherence should be rewarded (e.g., by congratulations messages), yet competition (e.g., leader boards) was not well received; patients preferred more private social experience, with a small group of friends and family [151].
  - Novel ways that provide patients with social support via the app should be developed and validated. Social support is not included in most existing apps.
b. Ginossar et al. presented a set of recommendations related to the usability of mHealth apps for breast cancer patients and the language used in these apps [150] that are applicable to other types of cancer:
  i. Usability recommendations include easy access to home page and clearly labeled back button, in-app simple search and browsing of provided content and use of images to facilitate learning
  ii. Language recommendations include using the pronoun “you”, present tense, active voice and action words, limiting the length of sentences and paragraphs, and using common everyday words and defining technical terms
c. According to Prochaska et al. [148] participation in content creation results in a stronger intention for behavior change (e.g. quitting smoking). Moreover, posting photos is associated with stronger engagement than for other types of posted materials.

It is interesting to note that the aspects of behavior most frequently considered when providing coaching include physical activity/sedentary behavior (also combined with diet), distress and anxiety, and self-efficacy.

#### 4.1.8 Coaching Systems Open Challenges

i. App design should rely on sound behavioral theories or models. Many available apps are intuitive and use formally defined digital interventions (e.g., captured by the initial taxonomy by Abraham and Michie [109] and by taxonomies revised for cancer apps [108]), and the frequently used categories of digital interventions involve: customization, information/behavior relationship, intention, facilitation, self-efficacy and social influence. However, identification of applied interventions is often performed only during post-hoc analysis, instead of being part of the design process.
ii. A controlled study should be used to measure improvement in QoL. Yet attributing improvement to particular system functionality necessitates large cohorts partitioned into groups receiving different app versions or a study in which different features are exposed over time, or even several studies (e.g., as for the CHESS system where different trials were conducted to assess the impact of several combinations of patient-oriented support [152]).
iii. Coaching systems for the palliative stage could potentially be developed; none of the papers in our review addressed this important stage, although home caregivers are likely to benefit from such systems.

### 4.3 Limitations

Our review study presents some limitations. First of all, the results presented in Section 3, and the discussion of best-practices and open research challenges presented in Section 4.1 are based solely on our particular set of papers reviewed.

A second limitation is that we did not include “well-being”, an important construct in cancer outcomes research, as part of our search keywords. However this was mitigated by the fact that the multidimensionality of QoL refers to a broad range of content, including the concepts of well-being [18].

## 5. Conclusion

We reported results of a systematic literature review about the state-of-the-art of computerized systems that employ Data Science methods to monitor the health status and provide support to cancer patients. Our analysis identified best practices that we intend to adopt in the CAPABLE project. In addition, we plan to address some of the open research challenges that are not currently addressed. These include supporting emotional and social dimensions of well-being, unobtrusive monitoring through wearable sensors, including PROs in predictive modeling and providing better customization of behavioral interventions for the specific population of cancer patients.

## Supporting information

CASP checklist

## Data Availability

No original data was used in the article

## Acknowledgements

The work described in this article has been funded by the European Union’s Horizon 2020 research and innovation programme under grant agreement No 875052 - CAPABLE (www.capable-project.eu).

