## Supplementary material for "A Review of AI and Data Science Support for Cancer Management": Table S1

Supplementary Table s1 - PubMed search strategy

| **Sub-topic** | **Search string** | **Last run** | **# papers** |
| --- | --- | --- | --- |
| Data collection | *cancer[tw]*  *AND (system[tw] OR app[tw] OR application[tw])*  *AND*  *(*  *(*  *(reporting[tw] OR monitoring[tw])*  *AND (*  *(QoL[tw] OR "quality of life"[tw])*  *OR (patient[tw] AND reported[tw] AND outcomes[tw])*  *)*  *)*  *OR*  *(*  *(monitoring[tw] OR sensors[tw])*  *AND patient[tw]*  *AND home[tw]*  *)*  *)* | 9/3/2020 | 318 |
| Data integration | *cancer[tw]*  *AND*  *("data integration"[tw] OR*  *(data[tw] AND (standards[tw] OR standard[tw] OR ontology[tw]) AND integration[tw])*  *)*  *AND (system[tw] OR app[tw] OR application[tw])* | 9/3/2020 | 136 |
| AI/prediction | *cancer[tw] prediction[tw]*  *AND*  *(machine learning OR AI OR "artificial intelligence")*  *AND*  *(stage OR state OR "quality of life")* | 9/3/2020 | 292 |
| Coaching systems | *cancer[tw]*  *AND (system[tw] OR app[tw] OR application[tw])*  *AND (*  *(coach[TW] OR coaching[tw])*  *OR ("behavior change"[tw] OR "behavioral change"[tw])*  *)* | 15/3/2020 | 85 |
